# “Safety and Efficacy of Exercise-based cardiac rehabilitation in patients with refractory angina: a randomized, controlled trial”

**DOI:** 10.1101/2024.08.08.24311708

**Authors:** Luciana Oliveira Cascaes Dourado, Camila Paixão Jordão, Marcelo Luiz Campos Vieira, Luis Henrique Wolff Gowdak, Carlos Eduardo Negrão, Luciana Diniz Nagem Janot de Matos

## Abstract

**Aim:** Evidence is scarce regarding safety and anti-ischemic effects of exercise-based cardiac rehabilitation (ECR) in patients with refractory angina (RA).

**Methods:** This was a prospective, single-center, randomized controlled study that assessed a 12-week ECR program in patients with RA. Death and cardiovascular events, anginal symptoms, exercise stress echocardiogram (ESE) and cardiopulmonary exercise test (CPET) parameters were evaluated. When significant differences were detected, Bonferroni post hoc comparisons were conducted.

**Results:** No difference regarding clinical events and anginal symptoms was found between groups. In ESE, rehab group (RG) increased in peak load (RG*post* to RG*pre*, *P* = 0.001; interaction, *P* < 0.001), angina quantification (RG*post* to RG*pre*, *P* = 0.004; control group (CG)*pre* than CG*post*, *P* = 0.006; RG*post* than CG*pre*, *P* = 0.0019; interaction, *P* = 0.001), exercise duration (RG*post* to RG*pre*, *P* = 0.009; interaction, *P* = 0.006), ischemic threshold (RG*post* to RG*pre*, *P* = 0.001; CG*pre* than CG*post*, *P* = 0.03; RG*post* than CG*pre*, *P* = 0.008; interaction, *P* = 0.005) and angina threshold (RG*post* to RG*pre*, *P* = 0.04; RG*post* than CG*post*, *P* = 0.04; interaction, *P* = 0.002). In CPET, RG had increased exercise duration and covered distance in RG*post* (RG*pre* than RG*post*, *P* = 0.001; interaction, *P* = 0.014, RG*pre* than RG*post*, *P* < 0.001; interaction, *P* < 0.01; respectively).

**Conclusion:** A 12-week ECR was safe and promoted positive clinical effects regarding exercise duration, intensity of angina, and angina and ischemic thresholds in RA patients.

The trial registry: Cardiac Rehabilitation in Patients with Refractory Angina (NCT03218891)

## INTRODUCTION

Refractory angina (RA) is a chronic and debilitating condition lasting >3 months, characterized by the presence of limiting angina due to myocardial ischemia in the setting of obstructive chronic coronary disease, not controlled by the combination of medical therapy, angioplasty and surgery.^1,2^

Patients suffering from RA experience significant quality of life impairment,^3^ and several promising alternative therapies, some experimental, have been tested with varying results.^4,5^ Therefore, it is urgent to incorporate safe, effective, and accessible therapies to minimize symptoms in RA.

Despite being a standard of care in chronic coronary artery disease (CAD) prevention and treatment,^6–8^ exercise-based cardiac rehabilitation (ECR) is not often prescribed to patients with RA due to scarce evidence supporting its safety and benefits in this scenario^9,10^ because these patients have a low ischemic threshold and substantial physical performance limitations.^11^ Therefore, these patients have been considered at high risk for exercise-induced adverse cardiac events.^6,8^

Moderate exercise training in patients with RA was previously shown to improve functional capacity in Progressive Shuttle Walk level attainment and angina threat perception, despite failing to improve quality of life perception.^9^ Even so, little is known about safety and anti-ischemic effects of ECR in patients with RA.

In this current study, we tested the hypothesis that 12-week ECR is a safe and effective strategy to improve anginal symptoms, physical performance, and stress echocardiographic ischemic burden in patients with RA.

## METHODS

### Study Design

This was a prospective, single-center, randomized controlled 12-week cardiac rehabilitation (CR) program in patients with RA conducted in a tertiary university hospital (São Paulo, Brazil). The study was approved by the ethics and research committee of the Hospital das Clínicas da Faculdade de Medicina da Universidade de São Paulo (CAAE: 24308213.7.0000.0068) and submitted to and approved by clinical trials.gov (NCT03218891). The study was conducted in compliance with the provisions of the Declaration of Helsinki. All patients provided written informed consent. The study was prematurely interrupted due to the COVID-19 pandemic.

### Study patients

Patients diagnosed with RA in clinical follow-up at a specialized outpatient clinic for RA in a tertiary university hospital were enrolled from April 2015 to January 2019. Candidates were of both sexes, aged 45 to 75 years, with symptomatic angina (CCS angina functional class from II to IV) >3 months of duration on optimal medical therapy (≥3 antianginal drugs). Patients included those who had myocardial ischemia that could be documented by physical stress echocardiography, and who were not eligible for surgical or percutaneous myocardial revascularization procedures.

### Exclusion criteria

Exclusion criteria were as follows: 1) permanent pacemakers or implantable cardiac defibrillators; 2) patients with non sinus rhythm; 3) history of recent (<3 months) acute coronary syndrome or myocardial revascularization (percutaneous or surgical); 4) functional impairment caused by any clinical condition preventing exercise; and 5) activity restriction (class D) according to the American Heart Association criteria for risk stratification of events during exercise.^12^

### Randomization and intervention

Patients meeting the inclusion criteria were randomized in blocks to ensure the sample homogeneity due to the sample size, with a computer-generated sequence to undergo either: 1) exercise-based cardiac rehabilitation group (RG), consisting of optimized clinical treatment + physical training for 12 weeks or 2) control group (CG), consisting of optimized medical treatment. Clinical and laboratory evaluations, cardiopulmonary exercise testing (CPET), and exercise stress echocardiography (ESE) were performed in all patients after inclusion and at the end of the study.

### Study endpoints

The prespecified endpoints were the evaluation of the safety of 12-week ECR in patients with RA, assessing clinical events (death and major cardiovascular events), in addition to the effects on anginal symptoms (angina functional class, angina attacks *per* week [AAW] and short-acting nitrate consumption *per* week [SANCW]), exercise stress echocardiography myocardial ischemic burden, and cardiopulmonary exercise testing physical performance.

### Clinical evaluation

Patients were evaluated during medical visits pre- and postintervention, consisting of a detailed medical history and clinical examination with anthropometric measurements. Angina functional class was evaluated according to Canadian Cardiovascular Society (CCS) classification.^13^ The AAW and SANC were recorded in an angina diary. Medical therapy adherence was encouraged during medical visits based on the patient’s tolerance. Antianginal drug prescription involved the combination of β-blockers, calcium channel blockers, short- and long-acting nitrates, trimetazidine and ivabradine, following the latest guidelines on the management of stable angina.^7^ Dosage of each medication was recorded as the percentage of the maximum recommended dose.

Laboratory exams were also collected at the same time points.

### Cardiopulmonary exercise testing

To verify effectiveness and to prescribe the exercise training protocol, CPET was performed pre- and post-intervention, on a motorized treadmill (T2100 Model, GE Healthcare, USA) and ergospirometer (SensorMedics – VmaxAnalyzer Assembly, Encore 29S, USA), using a graded exercise protocol (modified Balke 2.5 mph). Heart rate (HR) was continuously recorded using a 12-lead electrocardiogram (Ergo PC, Micromed, Brazil). Blood pressure (BP) was measured at rest, every two minutes of exercise, and on the 0th, 2nd, 4th, and 6th minutes of recovery. Oxygen uptake (VO_2_) and carbon dioxide production were determined by gas breath-by-breath exchange and reported an average of every 30 seconds. Maximal exercise capacity was determined by the VO_2_ measured at peak of exercise (VO_2_ peak) and also reported as a percent-predicted value using the Wasserman equation.^14^ Ventilatory threshold (VT1) was identified at the breakpoint between the increase in the carbon dioxide output and VO_2_ (V slope) or at the point in which the ventilatory equivalent for oxygen and end-tidal oxygen partial pressure curves reached their respective minimum values and began to rise.^11,15^ The angina threshold (AT) was determined at the exact time (in seconds) and HR at which the patient complained of angina symptoms. CPET was performed following the American Heart Association guidelines.^16^ Interruption was performed according to the III Brazilian Society of Cardiology Guidelines of the exercise test.^17^

### Exercise stress echocardiography (ESE)

To determine the ischemic burden, a two-dimensional echocardiogram evaluation was performed with the Vivid9 device (version 110.x.x, GE Healthcare) and according to the guidelines of the American Society of Echocardiography.^18^ After echocardiography at rest, exercise testing was performed on a lower limb cycle ergometer adapted to the stretcher, with a 45° inclination laterally and 45° horizontally. The workload was progressively increased from 5 to 25 watts every 3 minutes, according to physical capacity for each patient, and echocardiographic analyses were performed during all efforts. The exercise test was interrupted when patients reached exhaustion, limiting symptoms, the presence of significantly abnormal findings on the echocardiographic images, hemodynamic and/or electrocardiographic criteria according to guidelines.^17,18^ To assess left ventricular segmental contractility on ESE, an analysis 17-segmental model was used.^18^ Wall motion score index was defined based on American Society of Echocardiography guidelines.^18^ The onset of ischemia detected by ESE was determined at the time, and HR, which regional myocardial wall motion abnormalities (hypokinesis, akinesis, or dyskinesis) began, therefore considered the ischemic threshold (IT). AT was also recorded (HR and time). Angina intensity was graded according to a subjective scale of pain from 0 (no pain) to 10 (very intense).^19^

### Exercise-based cardiac rehabilitation

The ECR program was performed in a tertiary hospital cardiovascular rehabilitation center, inside a temperature-controlled training room equipped with cycles, treadmills, strength exercise equipment, free weights, mats, balls, and bands, and assisted by qualified rehabilitation professionals, including a physician and physical education professionals or physiotherapists. The protocol comprised 36 exercise-sessions, over 12 weeks (3 times a week sessions). Sixty-minute exercise sessions of supervised training were proposed: 40 minutes of aerobic training, 15 minutes of resistance training and 5 minutes of stretching. The aerobic exercise consisted of 5 minutes of warm-up, 30 minutes of continuous aerobic exercise on a motorized treadmill, according to the latest rehabilitation guidelines,^6^ and 5 minutes of cool-down. Aerobic exercise intensity prescription was guided by CPET, at a HR corresponding to at least VT1 and/or AT (in cases AT HR was below VT1 HR).

Continuous exercise was encouraged; however, if the patient experienced mild to moderate angina (up to 3), according to a scale graded from 0 (no pain) to 10 (severe pain), brief interruptions or reduction in intensity was recommended.^19^ Exercise was restarted when the symptoms were no longer observed. Patient was continuously monitored by telemetry. Sublingual isosorbide dinitrate (5mg) was administrated as needed. The resistance exercise session was based on the latest guidelines.^6,20^ Moderate resistance exercise intensity measured by perception exertion scale (level 4 to 6) was adopted^21^ and consisted of 2 sets of 8 to 15 repetitions. Resistance training was performed in a rhythmical manner, avoiding breath holding and straining (Valsalva maneuver) by exhaling during the contraction or exertion phase of the lift, and inhaling during the relaxation phase.^20^

### Statistical analysis and sample size calculation

Data were analyzed using SPSS Statistics 20 software. Continuous variables were expressed as mean ± standard deviation (SD) and categorical variables as percentages. The sample distribution was assessed using the Kolmogorov Smirnov test. For comparison of baseline characteristics between groups (RG and CG), the unpaired Student *t* test or Chi-square test was used, as appropriate. Generalized Estimating Equations (GEE) for longitudinal data analysis was performed. When significant differences were detected, Bonferroni post hoc comparisons were performed. Statistical significance was set at a *P* < 0.05. We designed a study with evaluation of continuous variables of independent intervention and control subjects. We considered the difference in sample response for each group of individuals to be normally distributed with a standard deviation of 1.2. If the mean difference among groups was 0.96, according to previous data^9^ evaluating functional capacity through the Shuttle Walking Test, we calculated the inclusion of 26 individuals for intervention and 26 individuals for the control group to ensure a minimum 80% power with a 5% significance level. The calculation of the sample size was performed using the PS program - Power and Sample Calculator version 3.0.43.

## RESULTS

The flow diagram (***Figure 1***) illustrates groups’ allocation, withdrawn and follow-up.

**Figure 1.**
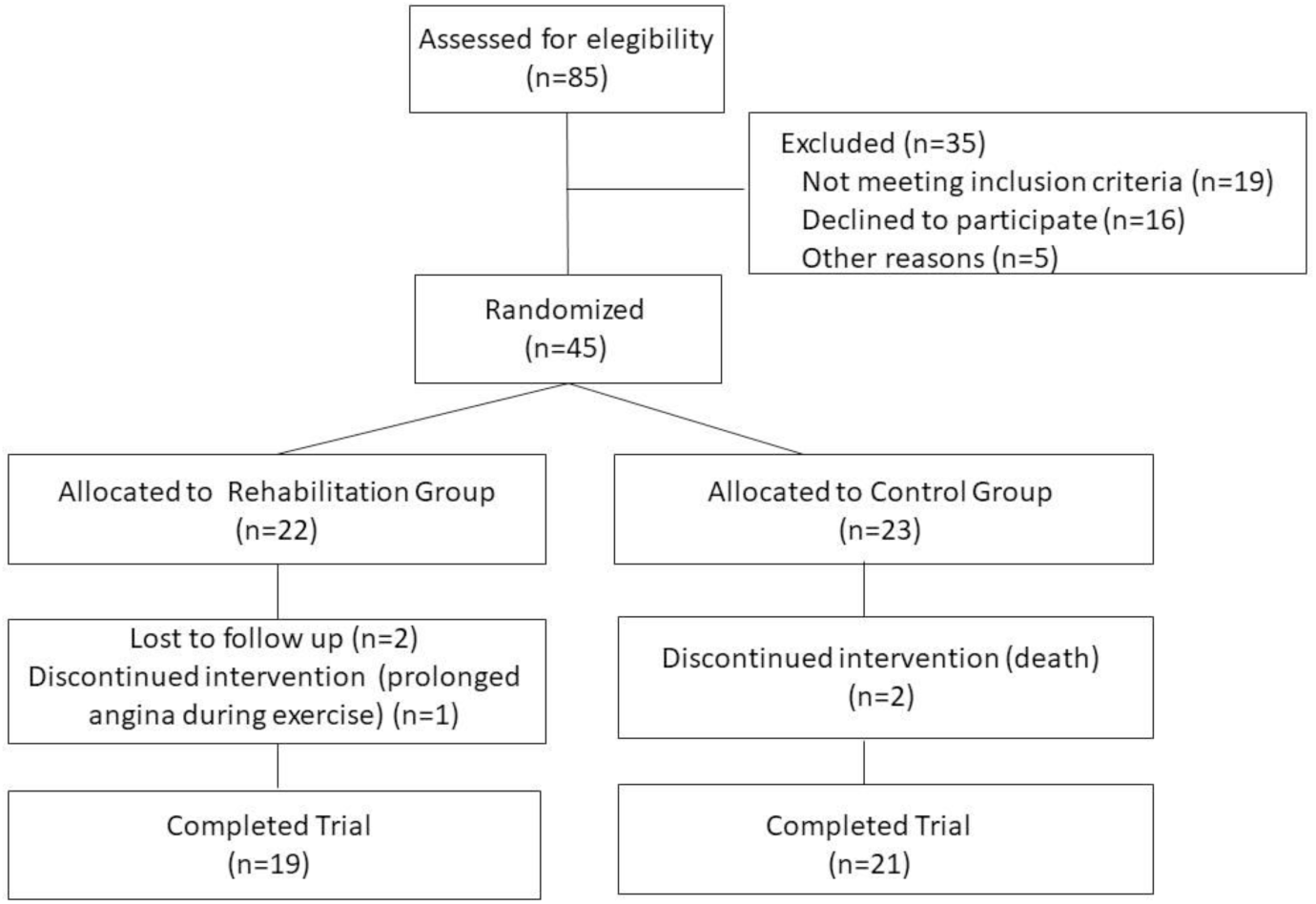
Flow diagram of rehabilitation group *versus* control group.

### Safety evaluation

Two patients in CG died during the study (1 cardiovascular death). One patient in RG experienced prolonged angina during a training session and was admitted to the emergency department for observation and was withdrawn from the study. There was no difference in major cardiovascular events between groups at the end of the study (***Table 1***).

**Table 1.**
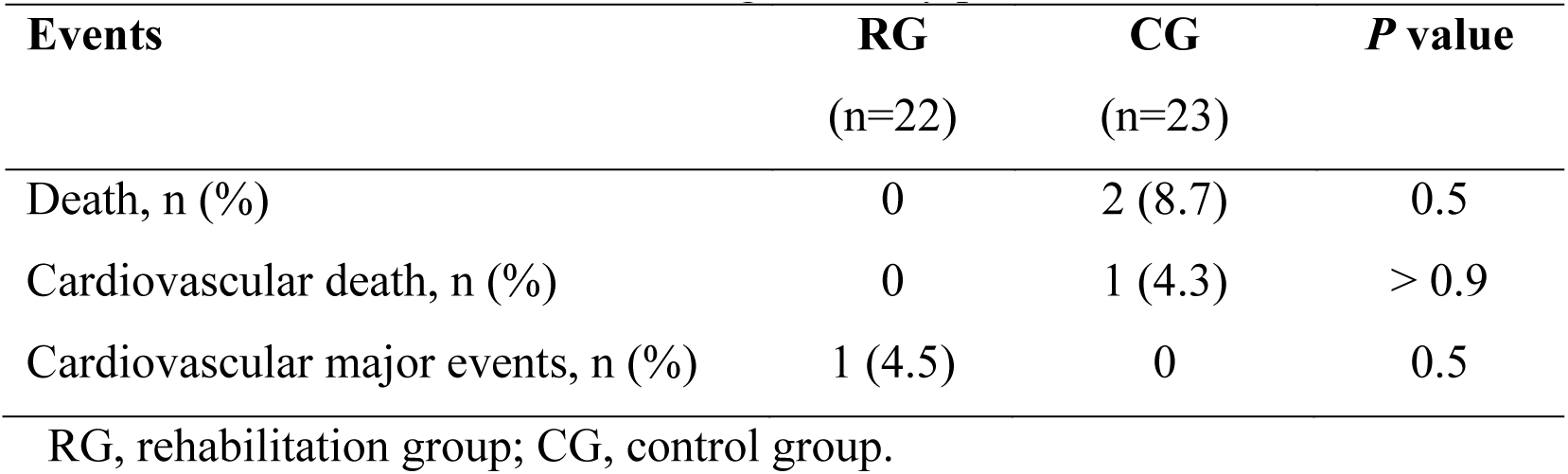
Cardiovascular events during the study period.

### Characteristics at baseline

The groups did not have significant clinical baseline differences (***Table 2***) regarding CCS angina functional class, AAW, SANCW, and clinical parameters. Patients of both groups had normal mean left ventricular ejection fraction (LVEF). Patients in RG have undergone more coronary interventions than those in CG. Complexity of the patients is demonstrated through high rates of risk factors, CAD anatomical pattern, coexisting conditions, and the high frequency of a combination of antianginal agents used (***Table 3***), in both groups. Laboratory findings did not identify significant differences except for HDL-cholesterol and triglycerides levels (***Table 2***).

**Table 2.**
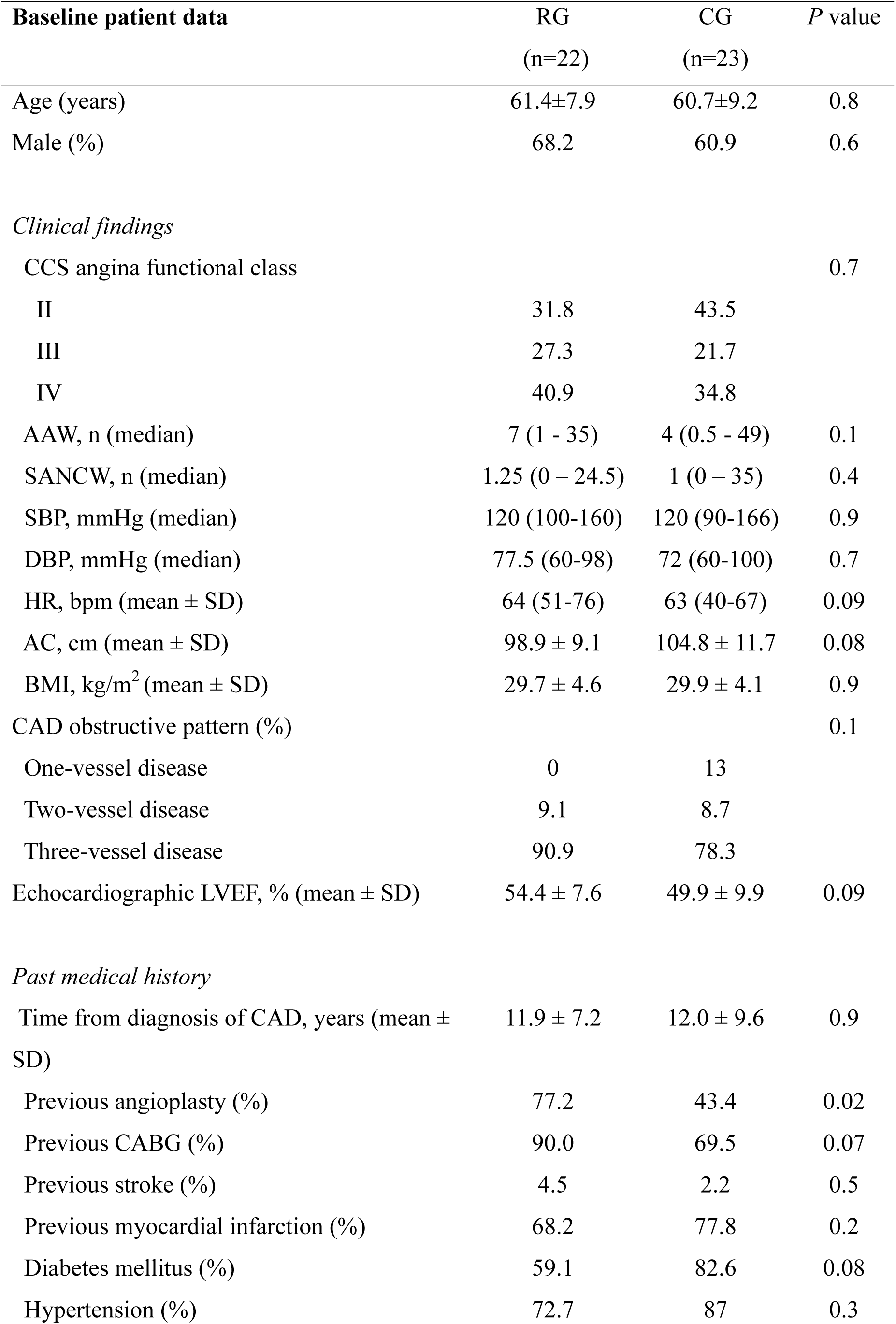
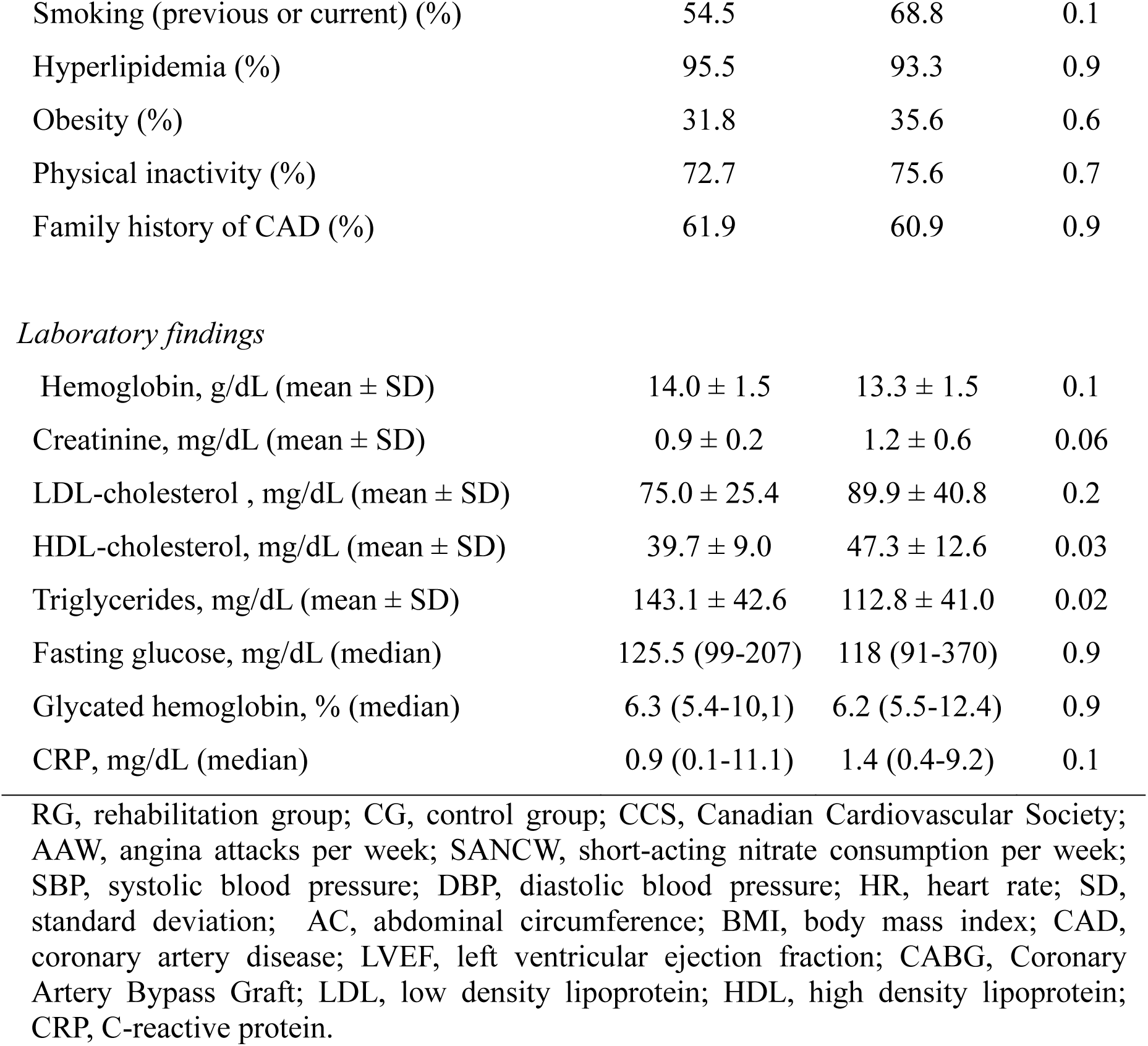
Demographic and clinical characteristics of patients at baseline.

| **Baseline patient data** | **RG** | **CG** | **P value** |
| --- | --- | --- | --- |
|  | **(n=22)** | **(n=23)** |  |
| Age (years) | 61.4±7.9 | 60.7±9.2 | 0.8 |
| Male (%) | 68.2 | 60.9 | 0.6 |
| *Clinical findings* |  |  |  |
| CCS angina functional class |  |  | 0.7 |
| II | 31.8 | 43.5 |  |
| III | 27.3 | 21.7 |  |
| IV | 40.9 | 34.8 |  |
| AAW, n (median) | 7 (1 - 35) | 4 (0.5 - 49) | 0.1 |
| SANCW, n (median) | 1.25 (0 – 24.5) | 1 (0 – 35) | 0.4 |
| SBP, mmHg (median) | 120 (100-160) | 120 (90-166) | 0.9 |
| DBP, mmHg (median) | 77.5 (60-98) | 72 (60-100) | 0.7 |
| HR, bpm (mean ± SD) | 64 (51-76) | 63 (40-67) | 0.09 |
| AC, cm (mean ± SD) | 98.9 ± 9.1 | 104.8 ± 11.7 | 0.08 |
| BMI, kg/m<sup>2</sup> (mean ± SD) | 29.7 ± 4.6 | 29.9 ± 4.1 | 0.9 |
| CAD obstructive pattern (%) |  |  | 0.1 |
| One-vessel disease | 0 | 13 |  |
| Two-vessel disease | 9.1 | 8.7 |  |
| Three-vessel disease | 90.9 | 78.3 |  |
| Echocardiographic LVEF, % (mean ± SD) | 54.4 ± 7.6 | 49.9 ± 9.9 | 0.09 |
| *Past medical history* |  |  |  |
| Time from diagnosis of CAD, years (mean ± SD) | 11.9 ± 7.2 | 12.0 ± 9.6 | 0.9 |
| Previous angioplasty (%) | 77.2 | 43.4 | 0.02 |
| Previous CABG (%) | 90.0 | 69.5 | 0.07 |
| Previous stroke (%) | 4.5 | 2.2 | 0.5 |
| Previous myocardial infarction (%) | 68.2 | 77.8 | 0.2 |
| Diabetes mellitus (%) | 59.1 | 82.6 | 0.08 |
| Hypertension (%) | 72.7 | 87 | 0.3 |
| Smoking (previous or current) (%) | 54.5 | 68.8 | 0.1 |
| Hyperlipidemia (%) | 95.5 | 93.3 | 0.9 |
| Obesity (%) | 31.8 | 35.6 | 0.6 |
| Physical inactivity (%) | 72.7 | 75.6 | 0.7 |
| Family history of CAD (%) | 61.9 | 60.9 | 0.9 |
| *Laboratory findings* |  |  |  |
| Hemoglobin, g/dL (mean ± SD) | 14.0 ± 1.5 | 13.3 ± 1.5 | 0.1 |
| Creatinine, mg/dL (mean ± SD) | 0.9 ± 0.2 | 1.2 ± 0.6 | 0.06 |
| LDL-cholesterol, mg/dL (mean ± SD) | 75.0 ± 25.4 | 89.9 ± 40.8 | 0.2 |
| HDL-cholesterol, mg/dL (mean ± SD) | 39.7 ± 9.0 | 47.3 ± 12.6 | 0.03 |
| Triglycerides, mg/dL (mean ± SD) | 143.1 ± 42.6 | 112.8 ± 41.0 | 0.02 |
| Fasting glucose, mg/dL (median) | 125.5 (99-207) | 118 (91-370) | 0.9 |
| Glycated hemoglobin, % (median) | 6.3 (5.4-10.1) | 6.2 (5.5-12.4) | 0.9 |
| CRP, mg/dL (median) | 0.9 (0.1-11.1) | 1.4 (0.4-9.2) | 0.1 |

RG, rehabilitation group; CG, control group; CCS, Canadian Cardiovascular Society; AAW, angina attacks per week; SANCW, short-acting nitrate consumption per week; SBP, systolic blood pressure; DBP, diastolic blood pressure; HR, heart rate; SD, standard deviation; AC, abdominal circumference; BMI, body mass index; CAD, coronary artery disease; LVEF, left ventricular ejection fraction; CABG, Coronary Artery Bypass Graft; LDL, low density lipoprotein; HDL, high density lipoprotein; CRP, C-reactive protein.

**Table 3.** Baseline drugs used by patients allocated in each group.

| **Medication** | **RG**<br>**(n = 22)** | **CG**<br>**(n = 23)** | **P value** |
| --- | --- | --- | --- |
| Aspirin |  |  |  |
| Patients, % | 95.5 | 95.7 | >0.9 |
| Clopidogrel |  |  |  |
| Patients, % | 40.9 | 17.4 | 0.08 |
| Statins |  |  |  |
| Patients, % | 100 | 100 | 1.0 |
| Ezetimibe |  |  |  |
| Patients, % | 9.1 | 17.4 | 0.7 |
| β-Blockers |  |  |  |
| Patients, % | 95.5 | 100 | 0.5 |
| % maximum dosage, median | 100 (25-100) | 75 (12,5-100) | 0.6 |
| Calcium channel blockers |  |  |  |
| Patients, % | 90.9 | 87 | >0.9 |
| % maximum dosage, median | 100 (25-100) | 100 (50-100) |  |
| Long-acting nitrates |  |  |  |
| Patients, % | 90.9 | 95.7 | 0.6 |
| % maximum dosage, median | 100 (50-100) | 100 (50-100) | 0.3 |
| Trimetazidine |  |  |  |
| Patients, % | 95.5 | 95.7 | >0.9 |
| Ivabradine |  |  |  |
| Patients, % | 81.8 | 87 | 0.7 |
| ACE inhibitors |  |  |  |
| Patients, % | 40.9 | 60.9 | 0.2 |
| % maximum dosage, median | 50 (25-100) | 100 (33-100) | 0.3 |
| ARBs |  |  |  |
| Patients, % | 22.7 | 30.4 | 0.6 |
| % maximum dosage, median | 100 (50-100) | 100 (50-100) | 0.4 |
| Diuretics |  |  |  |
| Patients, % | 36.4 | 47.8 | 0.4 |

| **Medication** | **RG**<br>**(n = 22)** | **CG**<br>**(n = 23)** | ***P* value** |
| --- | --- | --- | --- |
| Oral antidiabetic agents |  |  |  |
| Patients, % | 50.0 | 52.2 | 0.9 |
| Insulin |  |  |  |
| Patients, % | 27.3 | 30.4 | 0.8 |

RG, rehabilitation group; CG, control group; ACE, angiotensin-converting enzyme; ARBs, angiotensin receptor blockers.

### Effects of exercise training

#### Exercise training

Nineteen RG patients were engaged until the end of the protocol, 2 patients withdrew for personal reasons and 1 for a cardiovascular event; 2 CG patients withdrew due to death. Adherence to exercise training occurred in 76.7 ± 19.0% of sessions. Mean duration of aerobic exercise training was 39.3 ± 3.7 minutes. Mean HR during aerobic training was 78.7 ± 5.4 bpm, corresponding to 50.1 ± 4.3% of the maximum age-related HR. Angina HR during aerobic training was 81.3 ± 8.9 bpm, which was near the CPET angina/ischemic threshold. Fifteen patients (73.7%) experienced angina, and 5 of them (26.3%) needed to use a sublingual nitrate during at least one aerobic exercise training session. Mean systolic blood pressure and diastolic blood pressure during aerobic exercising was 121.7 ± 14.1 mmHg and 70.8 ± 5.6 mmHg, respectively.

No arrhythmias or ST-segment depressions occurred during the aerobic exercise training. Regarding resistance training, patients did not experience angina symptoms and/or any muscle discomfort; thus, resistance training occurred as expected.

### Clinical evaluation

After 12 weeks on protocol, there was no significant difference between groups regarding CCS angina functional class, AAW, SANCW, or clinical and laboratory parameters (***Table 4***).

**Table 4.** Clinical parameters changes in each group.

| Variable | RG |  | CG |  | P value |  |  |
| --- | --- | --- | --- | --- | --- | --- | --- |
|  | Pre | Post | Pre | Post | G | M | Int. |
| **Clinical data** |  |  |  |  |  |  |  |
| Angina functional class CCS, n (%) |  |  |  |  | 0.81 | 0.01 | 0.26 |
| 0 |  | 1 (5.3) |  |  |  |  |  |
| I |  | 6 (31.6) |  | 3 (14.3) |  |  |  |
| II | 7 (31.8) | 6 (31.6) | 10 (43.5) | 9 (42.9) |  |  |  |
| III | 6 (27.3) | 1 (5.3) | 5 (21.7) | 3 (14.3) |  |  |  |
| IV | 9 (40.9) | 5 (26.3) | 8 (34.8) | 6 (28.6) |  |  |  |
| Angina frequency. n | 11.6±10.1 | 7.18±12.5 | 8.8±11.1 | 10.4±11.1 | 0.76 | 0.26 | 0.05 |
| Nitrate consumption. n | 4.6±6.5 | 4.5±6.9 | 4.7±7.7 | 5.8±9.1 | 0.76 | 0.51 | 0.55 |
| SBP. mmHg | 124±16.8 | 119±16.5 | 123±17.6 | 121±20.2 | 0.87 | 0.26 | 0.70 |
| DBP. mmHg | 76.3±9.4 | 72.3±9.6 | 74.9±12.0 | 72.5±12.7 | 0.83 | 0.10 | 0.68 |
| HR. bpm | 63.3±6.5 | 61.7±5.7 | 59.3±7.4 | 63.3±9.6 | 0.51 | 0.25 | 0.02 |
| BMI. kg/m2 | 29.7±4.6 | 29.8±5.4 | 29.9±4.1 | 28.7±4.2 | 0.71 | 0.78 | 0.31 |
| **Laboratory data** |  |  |  |  |  |  |  |
| Fasting glucose | 131±31.5 | 123±26.8 | 149±72.8 | 143±82.8 | 0.22 | 0.53 | 0.66 |
| HBA1C. % | 6.7±1.3 | 6.5±0.9 | 7.0±1.8 | 6.8±1.6 | 0.35 | 0.76 | 0.37 |
| LDL-c | 75.1±25.4 | 72.8±23.2 | 89.9±40.9 | 93.2±42.9 | 0.145 | 0.56 | 0.94 |
| HDL-c | 39.7±9.0 | 40.0±9.9 | 47.3±12.7 | 48.1±10.3 | 0.033 | 0.79 | 0.25 |
| TG mL/g | 143±42.6 | 136±50 | 113±41 | 141±71.1 | 0.331 | 0.18 | 0.01 |
| CRP | 1.68±2.3 | 1.8±2.7 | 2.7±2.6 | 2.1±2.0 | 0.355 | 0.86 | 0.42 |

RG, rehabilitation group; CG, control group; G, group; M, moment; Int., interaction; CCS, Canadian Cardiovascular Society; SBP, systolic blood pressure; DBP, diastolic blood pressure; HR, heart rate; BMI, body mass index; HBA1C, glycosylated hemoglobin; LDL, low density lipoprotein; HDL, high density lipoprotein; CRP, C-reactive protein. Data are expressed as mean ± standard deviation or number (%).

### ESE evaluation

There was a significant increase in peak load (RG post to RG pre, *P* = 0.001; interaction, *P* < 0.001), angina quantification (RG post to RG pre, *P* = 0.004; CG pre to CG post, *P* = 0.006; RG post to CG pre, *P* = 0.0019; interaction, *P* = 0.001) (***Figure 2***), exercise duration (RG post to RG pre, *P* = 0.009; interaction, *P* = 0.006), time to ischemic threshold (RG post to RG pre, *P* = 0.001; CG pre to CG post, *P* = 0.03; RG post to CG pre, *P* = 0.008; interaction, *P* = 0.005) and time to angina threshold (RG post to RG pre, *P* = 0.04; RG post to CG post, *P* = 0.04; interaction, *P* = 0.002) between groups, as shown in ***Table 5*** and ***Figure 1 B, C, and D***, respectively. There were no differences in mean LVEF, ischemic burden score, peak HR, IT and AT HR (***Table 5***).

**Table 5.** Echocardiographic and cardiopulmonary parameters changes in each group.

| Variable | RG |  | CG |  | P value |  |  |
| --- | --- | --- | --- | --- | --- | --- | --- |
|  | Pre | Post | Pre | Post | G | M | Int. |
| **ESE** |  |  |  |  |  |  |  |
| LVEF, % | 54.5±7.6 | 53±7.7 | 50±9.9 | 48.8±9.9 | 0.06 | 0.03 | 0.67 |
| Rest score, n | 1.28±0.22 | 1.28±0.16 | 1.45±0.37 | 1.37±0.28 | 0.06 | 0.51 | 0.35 |
| Exercise score, n | 1.49±0.28 | 1.36±0.21 | 1.62±0.38 | 1.49±0.29 | 0.08 | <0.01 | 0.62 |
| Delta score, au | 0.21±0.16 | 0.08±0.11 | 0.16±0.11 | 0.12±0.11 | 0.95 | <0.01 | 0.10 |
| Exercise Duration, s | 291±89.5 | 344±143 | 277±110 | 266±93.7 | 0.07 | 0.02 | <0.01 |
| Peak load, watts | 34.7±17.8 | 46.2±23.8 | 35.2±24.2 | 35±21.4 | 0.31 | <0.01 | <0.01 |
| Angina quantification, n | 5.2±3.2 | 4.4±2.6 | 6.4±2.3 | 6.6±2.5 | 0.01 | 0.04 | <0.01 |
| Peak (HR), bpm | 96.1±9.7 | 95.5±18 | 99.5±15.8 | 94.8±11.9 | 0.84 | 0.58 | 0.07 |
| IT (HR), bpm | 90.5±7.2 | 93.5±12.7 | 93.6±15.7 | 91.6±10.5 | 0.93 | 0.88 | 0.20 |
| IT time, s | 241±86.4 | 299±136 | 210±74.9 | 214±83.4 | 0.02 | <0.01 | <0.01 |
| AT (HR), bpm | 89.1±7.5 | 89.2±8.9 | 92.1±16.5 | 90.6±10.2 | 0.60 | 0.70 | 0.65 |
| AT (time), s | 188±80.6 | 265±160 | 204±95.2 | 161±69.8 | 0.09 | 0.12 | <0.01 |
| **CPET** |  |  |  |  |  |  |  |
| VO<sub>2</sub> peak, mL.kg.min<sup>-1</sup> | 15.9±3.3 | 16.3±4.3 | 15.7±3.6 | 15.6±3.5 | 0.51 | 0.72 | 0.46 |
| VO<sub>2</sub> predicted, % | 63.8±19.2 | 64.5±18.3 | 60.2±14.8 | 61.3±10.3 | 0.31 | 0.80 | 0.78 |
| Angina quantification, n | 6.4±1.5 | 6.5±1.8 | 6.2±2.0 | 6.4±2.7 | 0.95 | 0.85 | 0.66 |
| VT1 (HR), bpm | 82.1±9.6 | 82.7±9.7 | 87.6±16.0 | 83.6±10.5 | 0.34 | 0.45 | 0.37 |
| AT (time), s | 198±66 | 322±174 | 248±132 | 287±94 | 0.86 | <0,05 | 0.07 |
| AT (HR), bpm | 85.9±13.9 | 88±13.1 | 92.5±16.3 | 90.7±15.2 | 0.33 | 0.95 | 0.36 |
| RQ | 0.9±0.99 | 1.0±0.1 | 1.0±0.1 | 1.0±0.1 | 0,93 | 0,74 | 0.01 |
| Oxygen pulse, ml/beat | 13.6±3,7 | 14.7±3.4 | 12,3±3,1 | 12.9±3.6 | 0.18 | <0.01 | 0.43 |
| Covered Distance, ms | 312±151 | 443±205 | 360±155 | 363±130 | 0.76 | <0.01 | <0.01 |
| Total Duration, s | 345±124 | 435±181 | 327±141 | 371±128 | 0.18 | <0.01 | <0.01 |

RG, rehabilitation group; CG, control group; G, group; M, moment; Int., interaction; ESE, exercise stress echocardiography; LVEF, left ventricular ejection fraction; HR, heart rate; IT, ischemic threshold; AT, angina threshold; CPET, cardiopulmonary exercise testing; VT1, Ventilatory threshold; RQ, respiratory quotient.

**Figure 2.**
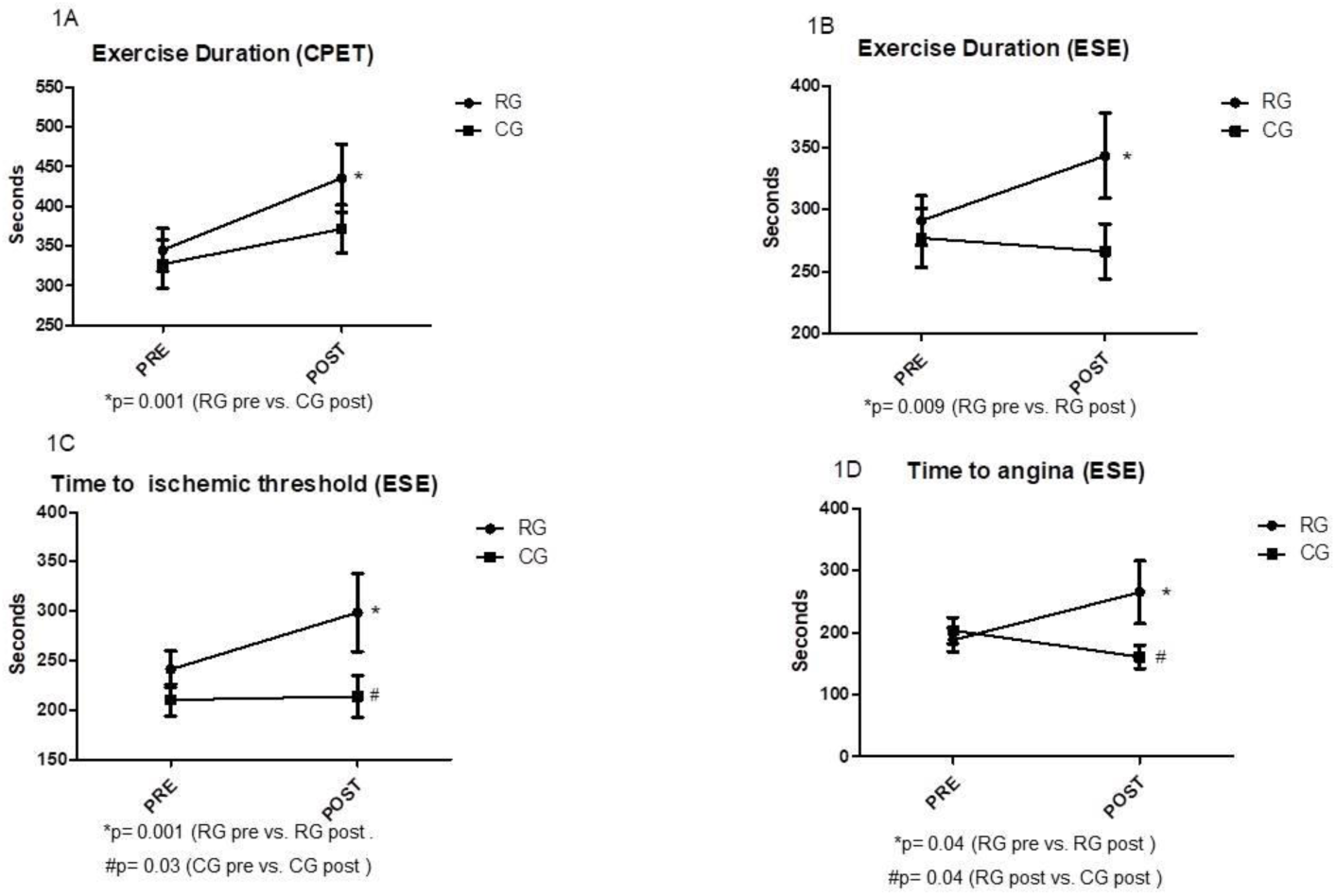
Change in exercise duration in cardiopulmonary exercise test, exercise duration, time to ischemic threshold and time to angina threshold in exercise stress echocardiogram, according to study group.

### CPET performance evaluation

There was a significant increase in CPET exercise duration and distance covered in RG post (RGpre to RGpost, *P* = 0.001; interaction, *P* = 0.014, RGpre to RGpost, *P* < 0.001; interaction, *P* < 0.01; respectively), as demonstrated in ***Table 5*** and ***Figure 1A***. No significant differences in VO_2_ peak and other CPET variables were observed (***Table 5***).

## DISCUSSION

For the first time, safety and anti-ischemic effects of ECR were objectively evaluated in patients with RA. Our results demonstrate that a 12-week ECR performed next to AT and/or VT1 was safe and had positive effects regarding exercise duration, angina intensity, ESE ischemia, and angina onset.

Since the first documentation of RA as a clinical problem in clinical practice, ECR has been a treatment option.^1,6,22^ Despite this, no strong evidence of safety exists that has supported its indication in the RA scenario and the how to apply it.^6,9,10,22^

In our study, patients experienced debilitating angina due to myocardial ischemia despite using, at least, 3 classes of antianginal agents, not being eligible for coronary intervention due to the complexity of coronary obstructive disease. Therefore, these patients had a high-risk profile for exercise-induced adverse cardiac events,^8^ and due to physical limitations, prescription - and safety execution - of the exercise was challenging, especially to achieve the target moderate intensity exercise.

On the other hand, exercising these patients, which prescriptions were guided by CPET parameters and angina threshold – indeed allowing the experience of angina up to 3 in a 10 scale, did not increase the risk of cardiovascular events during the study period. These data are reinforced by our previous data, demonstrating that an acute aerobic exercise session does not alter hs-cTnT in patients with RA, suggesting that no significant myocardial injury was elicited by exercising according to our suggested prescription.^10^

Despite failing to demonstrate improvement in clinical parameters referred by patients, as angina functional class (CCS), AAW and SANCW, we considered the improvement in exercise test parameters an important hallmark of ECR’s positive effect, because the increase in exercise time, reduction in angina intensity and improvement in the onset of angina can positively impact quality of life, which is the focus of RA management.

No significant improvement occurred in VO_2_peak measure. Furthermore, because of physical limitations due to the low angina threshold, keeping the target moderate intensity aerobic exercise was a challenge – sometimes unaffordable - and could explain the failure to improve VO_2_peak. Despite of this, even not increasing the VO peak (the most important prognostic CPET parameter), the increase in the distance performed in exercise testing can be interpreted as a clinical improvement, once it is also considered a prognostic marker in patients with heart disease.^23^

Therefore, RC is a feasible adjuvant therapy for patients with RA, supporting its indication. We believe that 12-week ECR is a starting point to exercise benefits in RA patients and sustained ECR may promote additional positive effects and improve quality of life.

### Limitations

The results of this analysis must be interpreted within the context of potential limitations.

1. The sample size calculation was based on the only previous study that evaluated the effect of cardiac rehabilitation in patients with refractory angina, despite different pre-determined outcomes.
2. The study was prematurely stopped due to the COVID-19 pandemic, including 88% of the calculated sample.
3. The strict inclusion criteria reflect the difficulty in selecting patients with refractory angina.

## CONCLUSION

A 12-week ECR was safe and promoted positive clinical effects regarding exercise duration, intensity of angina, and angina and ischemic thresholds in RA patients.

## Data Availability

The data that support the findings of this study are available from the corresponding author, LOCD, upon reasonable request.

## ACKNOWLEDGEMENTS

We are grateful to Camila R. A. de Assumpção and to Carla G. de S. P. Montenegro who have contributed to making this research possible.

## AUTHORS’ CONTRIBUTION

L.O.C.D, C.P.J.: conception, data acquisition, writing—original draft, and formal analysis. L.D.N.J.M: conception, writing—review & editing, and funding acquisition. M.L.C.V: data acquisition. L.H.W.G, C.E.N.: critically reviewed the manuscript. All authors read, gave final approval, and agreed to be accountable for all aspects of the work, ensuring its integrity.

